# The organisation of nurse staffing in intensive care units: a qualitative study

**DOI:** 10.1101/2022.01.18.22269459

**Authors:** Ruth Endacott, Chiara Dall’Ora, Annette Richardson, Peter Griffiths, Natalie Pattison, Susie Pearce, xSEISMIC research team

## Abstract

**Aims:** To examine the organisation of the nursing workforce in intensive care units and identify factors that influence how the workforce operates.

**Background:** Pre-pandemic UK survey data show that up to 60% of intensive care units (ICUs) did not meet locally agreed staffing numbers and 40% of ICUs were closing beds at least once a week because of workforce shortages, specifically nursing. Nurse staffing in ICUs is based on the assumption that sicker patients need more nursing resource than those recovering from critical illness. These standards are based on historical working, and expert professional consensus, deemed the weakest form of evidence.

**Methods:** Focus groups with health care professionals working in ICUs (n= 52 participants) and individual interviews with critical care network leads and policy leads (n= 14 participants) in England between December 2019 and July 2020. Data were analysed using framework analysis.

**Findings:** Three themes were identified: the constraining or enabling nature of ICU and hospital structures; whole team processes to mitigate nurse staffing shortfalls; and the impact of nurse staffing on patient, staff and ICU flow outcomes. Staff made decisions about staffing throughout a shift and were influenced by a combination of factors illuminated in the three themes.

**Conclusions:** Whilst nurse: patient ratios were clearly used to set the nursing establishment, it was clear that rostering and allocation/re-allocation during a shift took into account many other factors, such as patient and family nursing needs, staff wellbeing, ICU layout and the experience, and availability, of other members of the multi-professional team. This has important implications for future planning for ICU nurse staffing and highlights important factors to be accounted for in future research studies.

**Implications for Nursing Management:** The potential opportunities for different staffing models are likely to be highly dependent on other professions. Hence, any change to staffing models needs to take into account how different professions work together.

Factors such as ICU layout, reported to influence nurse staffing decisions, suggest that patient safety in ICU may not be best served by blanket ‘ratio’ approaches to nurse staffing, intended to apply uniformly across health services.

The findings have the potential to feed into discussions about funding tariffs for critical care and quality metrics to be included in commissioning contracts.

## Introduction

Pre-pandemic UK survey data show that up to 60% of intensive care units (ICUs) did not meet locally agreed staffing numbers and 40% of ICUs were closing beds at least once a week because of workforce shortages, specifically nursing (FICM, 2018). Nurse staffing in ICUs is based on the assumption that sicker patients need more nursing resource than those recovering from critical illness. These standards are based on historical working, and expert professional consensus, deemed the weakest form of evidence (Howick et al 2012). For a fuller description see Endacott et al (2021).

In England, critical care services are supported by 15 Critical Care Networks (NHS Commissioning Board, 2012). Critical care networks support the coordination of patient pathways between health services to ensure access to specialist support at a regional level and benchmark services to ensure consistency, including peer review (NHSE 2021). Critical care networks also monitor workforce levels and provide support to individual ICUs, particularly to nurse and medical critical care leads. It is timely to examine how ICU nurse staffing is organised in this resource-constrained climate and what factors influence how the workforce operates.

## Background

A recent systematic review of 55 studies found significant associations between low ICU nurse staffing and worse outcomes for patients, staff and health services (Rae et al 2021). Odds of nosocomial infection were 3.28-3.60 times higher in ICUs with lower levels of nurse staffing and mortality odds were 1.24-3.50 times greater (Rae et al 2021). West et al (2014) found that significant associations between number of nurses and mortality had the greatest impact on patients at higher risk of death. Rae et al (2021) highlighted the wide variation in the way nurse staffing is defined and organised. However, the most common approach was some form of ratio (nurse:patient or nurse:bed). From their systematic review of ICU nursing workload instruments, Greaves et al concluded that no workload instrument has been adequately validated for this purpose, or found to be superior to the clinical judgment of an experienced ICU nurse manager (Greaves et al 2018).

Persistent registered nurse and ICU-qualified nurse staffing challenges, with vacancy rates at 10% across the UK (CC3N, 2020), have led to consideration of alternative staffing models. Coupled with the development of new roles, such as critical care nursing associates (Bates, 2019), and physician assistant/assistant practitioner roles in ICU, the workforce operates in an increasingly diverse way across the UK. The evidence around these changes is, as yet, lacking but a need to better understand what influences how this new workforce is deployed has been articulated (Henshall et al 2019), not least as there are several implications around role boundaries that have a direct impact on staffing decisions.

## Aims

The aims of the study were to examine the organisation of the nursing workforce in intensive care units and identify factors that influence how the workforce operates.

### Design

A qualitative in-depth exploration of the organisation of nurse staffing in ICUs from the perspectives of multi-disciplinary ICU teams, critical care networks and policy leads. Focus Groups were conducted with a mix of uni-professional (nursing) and multi-professional teams in ICUs and individual interviews were conducted with network leaders and policy makers.

### Participants

Purposive sampling was used to recruit nurses and other members of the ICU multi-professional, critical care network leaders, national and regional policy makers to the study. The concept of *information power* (Malterud et al., 2015) underpinned sample size, the premise being that the larger the information power of the sample, the smaller the sample required to achieve saturation. It was anticipated that the network and policy participants would provide broad insight across a number of ICUs. Policy-makers were drawn from NHS England and Improvement, the critical care Clinical Reference Group and clinicians responsible for commissioning ICU services at regional and national level. Representing most regions in England, 14 individual interviews were conducted with network leaders and policy makers. All bar two of the interview participants had an ICU clinical background. The network participants were responsible for networks across England covering between eight and 21 ICUs, a total of 145 ICUs. Six Focus Group interviews, lasting on average just over 80 minutes, were conducted with 52 healthcare professionals (nurses, allied health professionals, doctors and nursing assistants) from four hospitals and eight ICUs across England. The number of participants at each focus group ranged from 4-12 and participants worked in general ICUs (n=4), cardiac ICUs (n=2) and neurosurgical ICUs (n=2) in hospitals of varying bed size and patient population. Professional roles for all 66 participants are provided at Table 1.

**Table 1.**
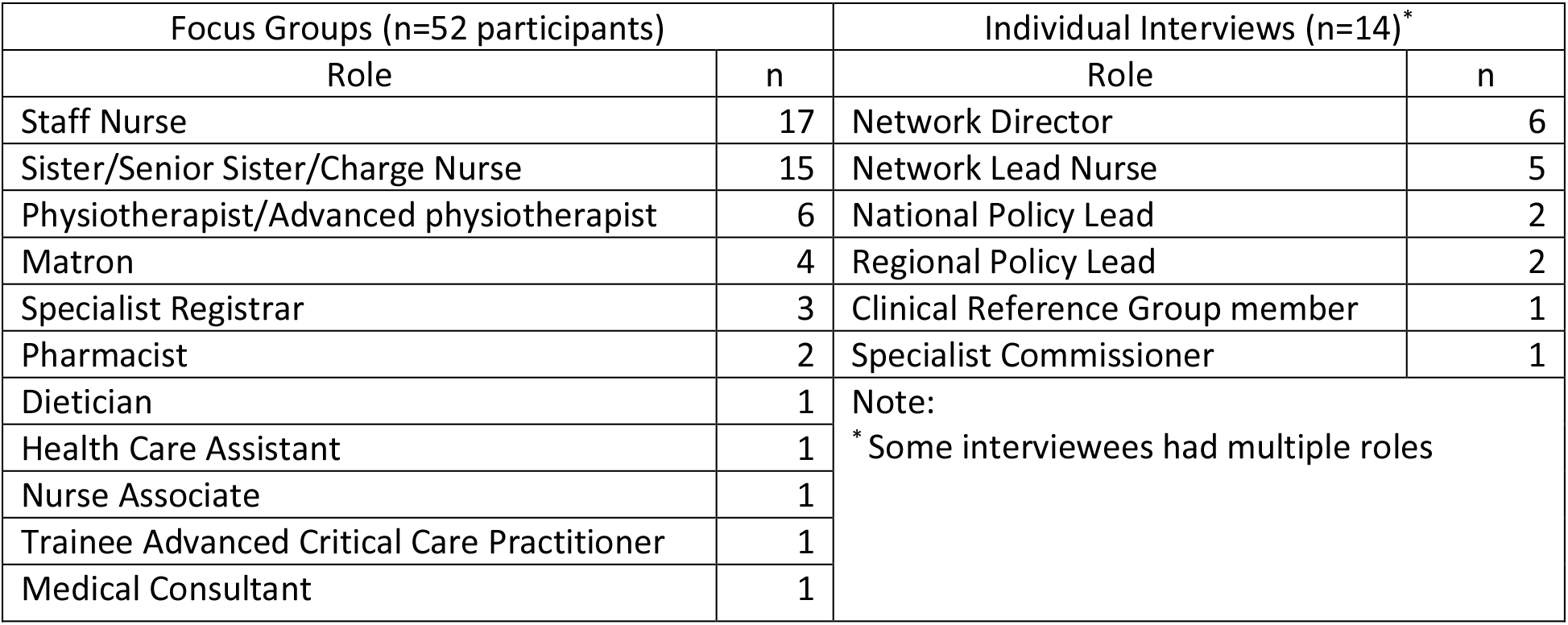
Participant roles.

### Data Collection

Study design and conduct were underpinned by principles of trustworthiness (credibility, transferability, dependability and confirmability) (Lincoln & Guba 1985). The interview topic guides were developed with an expert advisory group external to the research team (the UK Critical Care Nursing Alliance, which is the alliance uniting all critical care nursing organisations in the UK).

Individual interviews were conducted by RE and Focus Groups were led by SP with support from one other member of the study team. RE and SP are highly experienced qualitative researchers with extensive backgrounds in research related to the organisational aspects of care delivery. The interviewers (RE & SP) had regular meetings during data collection and analysis. Early analysis was reviewed by the whole research team and analytical memos were shared. To ensure a strong connection between the analysis and clinical perspectives, emerging themes were discussed with a clinical stakeholder group. Focus Groups were conducted at the health services from December 2019 to February 2020 (pre-pandemic), and the individual interviews were conducted online between July-September 2020, the early stages of the COVID-19 pandemic. The individual interviews had two components: pre-pandemic staffing (reported here) and the impact of COVID-19 on staffing models (reported in Endacott et al, in press).

### Data Analysis

Framework analysis was used, a process comprising five stages: familiarisation, defining a thematic framework, indexing, charting and mapping/ interpretation (Pope *et al*., 2000). The Focus Groups were conducted first and the individual interviews built on the initial themes. Early analysis identified that nurse staffing was commonly discussed in relation to safety, and nurse staffing was described in terms of structures, processes and outcomes. Hence, we used the Donabedian model of structures, processes and outcomes to structure the *a priori* framework for the analysis (Donabedian 1988); Donabedian’s model is one of the most dominant quality evaluation instruments used in healthcare and has been used in the ICU context, for example, to develop ICU patient safety indicators (Wu 2020).

### Ethical considerations

Participants were contacted prior to planned interviews and given sufficient time to consider participation. Audio-recorded verbal informed consent was sought prior to each interview. Focus Group participants provided written consent in person. Focus groups and interviews were audio-recorded and transcribed verbatim by a transcribing company. A favourable research ethics opinion was provided by Yorkshire and Humber Research Ethics Committee (REC ref no: 19/YH/0284) and the Health Research Authority (IRAS ID 259475).

### Findings

The categories initially used for the framework were: the staffing establishment, rosters, staff allocation structures and decisions, multi-professional teamwork, communication and outcomes for patients and staff. These were iteratively refined as coding and categorisation developed, in line with framework analysis methods (Gale et al 2019). The findings are reported using Donabedian’s model of structures, processes and outcomes. Three themes were identified: the constraining or enabling nature of structures; whole team processes to mitigate nurse staffing shortfalls; and the impact on patient, staff and ICU throughput outcomes. Further categories emerged during the analysis; the final framework with themes and categories is provided at Figure 1.

**Figure 1.**
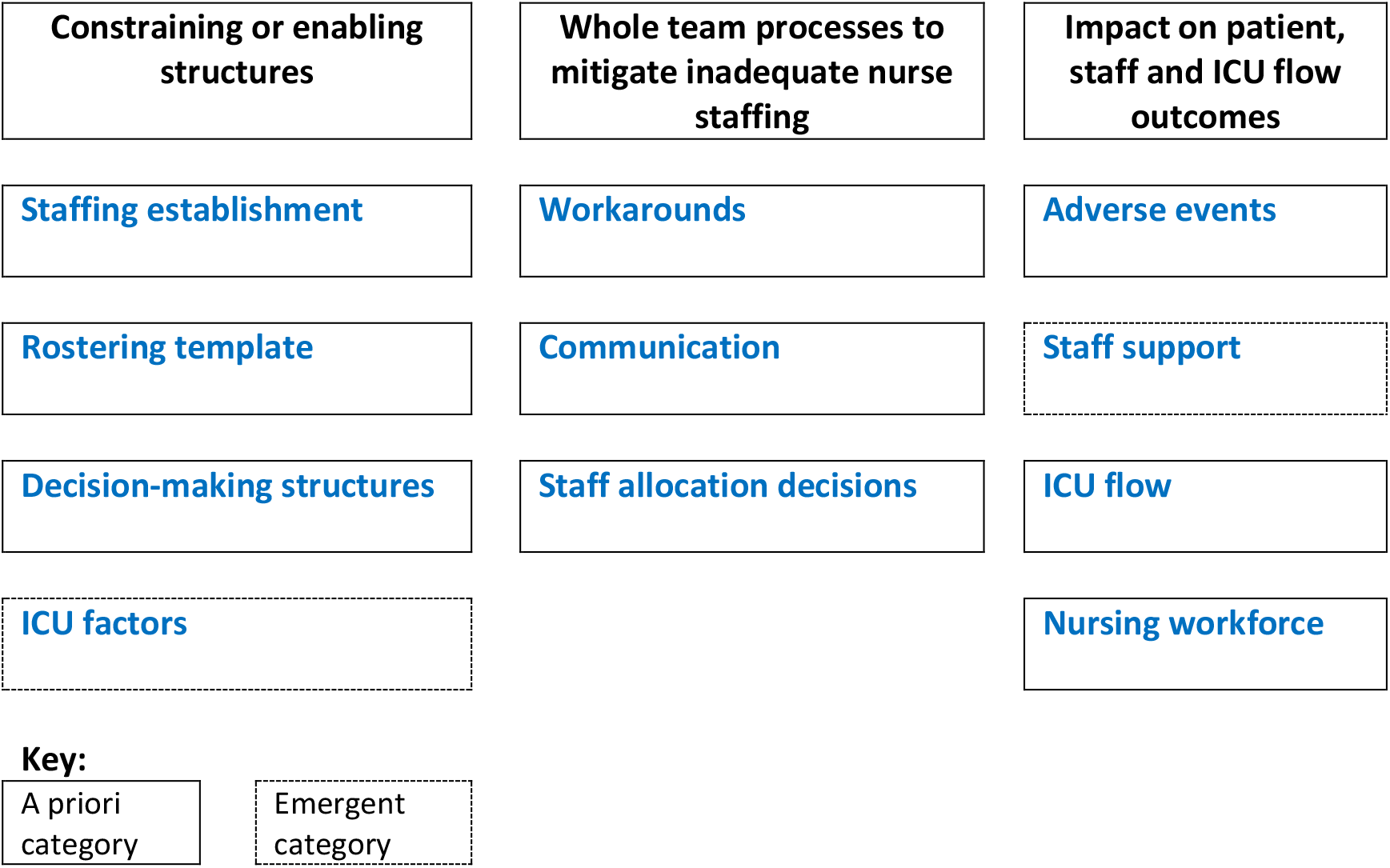
Managing nurse staffing: overview of themes and categories.

Data excerpts presented below and in tables are annotated as follows: *FG* denotes Focus Group and participant type, by profession; *Int* refers to interview and participant category (Network Lead or Policy Maker).

#### The constraining or enabling nature of structures

Many of the organisation-wide and ICU-specific structures described by participants related to the staffing models used to set the nursing establishment and the rostering template used to provide nurse staffing for each shift. The other categories in this theme are decision-making structures and ICU factors (see Table 2). These were all described in terms of the extent to which they enabled or constrained the organisation of ICU nurse staffing.

**Table 2.** Data excerpts related to *constraining or enabling structures* theme.

| **Structures** |  |
| --- | --- |
| Staffing establishment | *That's where an experienced group of people working together – and I'm not talking skill mix I'm talking just experience with – you know – team dynamics and the demands of the unit – if they work well together then it's easier to overcome the challenges that come along the way and everybody still gets breaks and drinks. Than when you've got junior staff (FG4/staff nurse)*<br>*Because that [reduced skill mix] just increases the pressure on you to micro-manage each individual bed space ...if you've got a whole load of new grads in an area then you're trying to go, 'Well, have you done that? Is the inotrope being reduced? Are you doing that?' and you can almost trust less in a way. (FG1/staff nurse)*<br>*So, we don't plough enough into building our staff's resilience and robustness.. We don't have a psychology service. Absolutely, desperately, need one. Trialled one a couple of years ago but then no more money got put into it. So, we know the need is there. (FG2/Matron)* |
| Rostering template | *I think we probably use our beds slightly differently but the staffing models are fairly traditional and well established... 1:1 and 1: 2 following the national guidance. (Int1/Policy Maker)*<br>*So, SafeCare system [rostering software] – you can add on tasks. Not all organisations do this... but we have added on our five person turns, for instance, and our side rooms. So, we have built that in trust-wide so that does tell us what the workload indicates and that's how many hours you're short in care hours. So, we are getting there.... We've got the data to show that there's a big gap between the number of staff hours we have and the number of care hours we have. (FG3/matron)*<br>*Our Nurse Dependency Scoring is fully embedded into the units and everyone uses it now. Based on our nurse dependency score, if they've got somebody who's a new admission, needs a lot of interventions and is really sick then they will bring in more staff to support that patient. I know a lot* |
|  | *of the senior nurses use it as a defence against finance coming on and wanting to cut your staff, and also to explain to doctors what the nursing workload really is, as opposed to what they think it is based on pure physiology [how sick the patient is].(FG8/Network Lead)* |
| Decision-making structures | *From a trust-wide perspective. We all meet. All together. Across every single care group at 11 o clock and we look at everyone's staffing and see which area can help others. (FG2/Matron)*<br>*When you don't have enough staff, you then escalate to agency, high cost agency, and that's a matron decision or Head of Nursing decision. (FG3/Matron)*<br>*We commission critical care and when critical care nurses are pulled off to go and work on the ward, that doesn't help with recruitment and retention. Also, we are paying those nurses to be on critical care and they're ending up elsewhere. So, we did put out a framework about what our expectations were about the use of critical care nurses and they shouldn't be routinely being pulled off critical care. (Int 13/Policy Lead)* |
| ICU factors | *We don't leave our patients alone so there's always a nurse next to them. So, if there is someone that is in a side room, someone has to come in, physically, and cover you to get medication, go to the toilet. For absolutely everything. (FG3/staff nurse)*<br>*if you're managing two cohorted critical care sites within one – what used to be one – critical care unit – that means you need more senior staff. You need more coordinators. Because you can't say, 'Well, that's the annexe so we're going to do that at a less standard and the patients in there are going to be compromised.' (FG8/Network Lead)*<br>*Sister: As the nurse in charge, I might end up with three level two patients but if one is in bed space 2, the other's in 10, the other's down in 14, they all require a nurse because the location does not allow me to split a nurse into three or into two safely .....*<br>*Staff Nurse Yeah ...*<br>*Staff Nurse.... and often they're in side rooms for a reason ...*<br>(FG1/Staff Nurse/Sister/Staff Nurse) |

Discussions about the staffing establishment focused on number of staff and the skill-mix required; there was evidently a complexity and interaction between roles. The inclusion of support staff (referred to by participants as Band 4, health care assistants (HCAs) or nurse associates) in the staffing establishment has been standard practice for a long time: ‘*we’ve had band 4 probably on our shop floor taking patients alongside a registered nurse for over 10 years’* (FG4/Sister). These roles were also deemed important: ‘*we didn’t used to have any but, in a way, it’s probably made more of a difference* (having an HCA) *than having another nurse*’ (FG5/Staff Nurse).

The skills of support (non-registered) staff were particularly valued in the care of long-term patients ‘*band 4* (senior) *HCAs have a more holistic view* [than junior support staff], *they’ve built a relationship with the family so they know the patient a bit better* ‘ (Int5/Network Lead) and patients with delirium (FG7/Staff Nurse). The importance of supernumerary roles (for example, shift leader, practice educator) was also emphasised, although they were also described as ‘*on the substitutes’ bench – when it’s busy they have to take a patient, which happened about 25% of the time over the past year* (Int 1/Policy Maker).

The number and skill mix of the multi-disciplinary team (MDT) also had an impact on the organisation of nurse staffing: ‘*also ‘how many physios are there? How many doctors? Is it the weekend?’ because there’s less* (other MDT members) *about then*’ (FG6/Matron). Doctors also acknowledged that:

> ‘(medical) *staff have become more junior so we can’t guarantee there’s always someone who can intubate and ventilate on the unit, meaning we rely on the advanced critical care practitioners* (senior ICU nurses)*’* (FG7/Doctor).

Staff rostering structures were discussed; all sites described some form of rostering template, grouping nursing staff according to skill-mix, to keep the staff and patients as safe as possible. However, hospital-wide quotas for annual leave, maternity leave and study leave also had an impact on rostering: ‘*to just get all the* [nursing] *staff to do their mandatory study time takes us over the quota we’re allowed, without allowing any time for ICU education’* (FG4/Matron).

Nurse:patient ratios, linked to the designation of individual patients as intensive care (level 3) or high dependency (level 2), did not seem to work for high dependency patients:

> *…you know – an intubated ventilated level three patient who’s not on a filter* (renal haemofiltration) *but just needs regular rolling and some early physio can actually quite often be easier to manage from a workload point of view than a level two delirious patient who’s taking three people to hold them in the bed and stop them ripping their lines out. There’s no – yeah – there’s no measure of those things*. (FG1/Sister)

There was a clear sense of a shift away from organ support as the rationale for nurse staffing ‘*we need 1:1 nurse patient ratios for different reasons now* (than when ratios were first introduced 20 years ago*)’* (Int 8/Network Lead). This was also referred to as needing ‘more columns’ in the

> *P1: …*..*the rehab patients are not an organ failure in the strictest sense of the word. They’re not on filtration* (renal support). *They’re on minimum amounts of oxygen but they are requiring the services of one nurse just to look after them in the way that they should be looked after*. (FG5/Doctor)

decision-making algorithm:

However, whilst it was suggested that ‘*most units don’t measure against the ratios, day to day*’ (Int 10/Network Lead), they were deemed to provide a safe standard that was easily communicated from junior ICU nurse right through to Board Members. As a policy lead (and ICU clinician) commented: ‘*you’re aiming for it or you’re not; you’re achieving it or you’re not… it allows a more coherent discussion about risk*’ (Int 1/Policy Lead). There were concerns that the loss of this clear standard would put ICU nurses in the same (unacceptable and understaffed) position as hospital ward nurses (Int 4/Network Lead). Nurse:patient ratios were also deemed valuable to develop a business case for staffing: ‘*it’s fairly clear how many nurses you need to open an extra critical care bed*’ (Int 1/Policy Lead).

The executive-level hospital decision-making structures were also perceived to be important; however, the inclusion of non-clinical managers in staffing decisions caused frustration and anger:

> ‘*as senior nurses we have lost the ability to manage our staff within intensive care …the key frustration is when it’s the Housekeeping Supervisor, who happens to be the on-call manager, telling you to move staff to the ward*’ (FG1/Sister)

Having ‘*a champion for ICU at executive level’* (Int 9/Network Lead) was deemed to make a positive difference, as well as twice-daily meetings to look at hospital-wide staffing (FG2, FG5). A longer-term impact of hospital staffing policies related to promotion criteria with nurses in some sites having to leave ICU to achieve promotion (Int 4, Int 8, FG4). This was also reflected in the nursing staff impact category, under the Outcomes theme (see Table 2).

Two issues emerged during the analysis: challenges in identifying staffing across hospitals and factors related to the individual ICU. There were some challenges in identifying exactly what the staffing is across different hospitals because of differences in coding: ‘*You think on an electronic staff record that we would all code the same but we don’t. So, you can’t at any point say exactly how many registered nurse vacancies you’ve got. Or how many registered nurses have you got* [across hospitals]. *Because a clinical nurse specialist might not be coded as a nurse’* (Int 1/Policy Lead).

Individual ICU factors such as visiting policies (FG6), admission policies (FG2), the size of the ICU (Int4) and the mix of emergency and elective patients admitted (FG7) influenced how nurse staffing was managed. The latter point had an important influence on processes, in particular the frequency of workarounds, articulated more frequently in ICUs with fewer elective patients. However, the layout of the ICU was the most frequently discussed factor across Focus Groups and individual interviews. This was important for structure and process factors as it affected whether the staffing establishment was correct for that particular ICU but also affected how staff were allocated and re-allocated during the shift (see Table 2).

At one health service, a new ICU layout, with mainly side rooms, resulted in all patients needing a 1:1 ratio because staff were in a side room and unable to oversee more than one patient due to the physical environment (Int 11/Network Lead). This led to an increased staffing establishment, particularly supernumerary staff, in order to manage breaks and provide adequate supervision; however, the nurse manager had to defend these decisions by developing ‘*a checklist of reasons why patients can’t be left alone in side rooms, to justify staffing* [to managers outside of ICU]’ (Int 11/Network Lead).

#### Workarounds: whole team processes to mitigate inadequate nurse staffing

The processes described were positive steps taken to continue care provision when staffing was challenging, often undertaken in rapidly changing situations, such as emergency admissions, patients deteriorating. The main categories were workarounds, communication and staff allocation decisions (see data excerpts at Table 3).

**Table 3.** Data excerpts related to *whole team processes to mitigate inadequate nurse staffing* theme.

| Processes |  |
| --- | --- |
| Workarounds | *The physios and the techs, they will muck in. They will come and help. They will do turns. They will help a lot if they feel – if we need them to. So, we have that extra support which helps. Consultants will help; they're very visible. They're very hands-on. (FG3/Matron)*<br>*If we need to get a patient in [when we short of nurses] ... then you need to present that to them: "We're at full stretch from a nursing point of view. I haven't got a nurse for this [patient] but bring them into our safe zone, which is in easy sight of everything and anything that you could need. But you [doctor] will have to remain with them". (FG5/Matron)* |
| Communication | *I see it then as well in also our interactions with people like nurse in charge because your concentration is far more divided on those days when you don't have a good skill mix...for things like ward rounds having the ability for the nurse in charge to come around and actually focus on the plans rather than jiggling staff and all that sort of thing. (FG4/Doctor)*<br>*[Which nurse is in the bed space] that makes a huge difference as a doctor coming in, where you only get snapshots of patients. The information you get fed from nursing staff and having either an experienced nurse overseeing or – you know – the right people in the bed space is key to getting that information. (FG1/Doctor)*<br>*So, you're getting the handover and you're thinking, 'What's gone on? And who's made that decision?' ....I think that's when I just turn around and I just go, 'Well, I feel unsafe doing this' but it's taken nearly six years [of experience in ICU] to say, 'I'm not happy. I don't think this is safe. Can we do something about it?' (FG3/staff nurse)* |
| Staff allocation decisions | *I mean there's nothing more frustrating than [when] you're about to have a really important conversation with the family, you really need the nurse to be there with you, but there isn't the staff to free up the nurse to come along... or it takes 10 minutes of jiggling to free somebody up...(FG1/doctor)*<br>*Sometimes it's not about the amount of courses they've done. Sometimes it's just about confidence and trust. Actually do I trust that nurse to keep an eye on my patient and know that they're gonna escalate if something went wrong? (FG3/matron)*<br>*What we do in [name of network] is that staff use flexi-staffing; instead of coming into the unit and being sent to the ward, they'll bank hours and they'll go home [and work those hours another time]... They prefer to do that, essentially unpaid on call, knowing that they could potentially get called back in...rather than be sent to a ward. (Int9/Network Lead)* |

The most dominant processes talked about across the Focus Groups and individual interviews were ‘work-arounds’. These were decisions and practices undertaken to reach a practicable solution. A picture was presented of a practised fluidity in the blurring of roles across professions when nurse staffing levels were inadequate, from the relatively short-term ‘*do you mind if I get something while you’re here* (staff nurse talking to physio)*?*’ (FG4/N24) to more substantive examples of physiotherapists informally doing nursing ‘work’:

> ***Physio 1:*** *We’ll do the obs*[ervations] *while people are on break…*
>
> ***GROUP:*** *(murmurs of agreement)*.
>
> ***Physio 1:*** *… that’s so the nursing staff can have a break, then nursing staff will help us with stuff. And all of a sudden you are a morphed, merged MDT that’s making the best of a bad situation…. Those days are more frequent than not…*. (FG1/Physio)

Communication with family members was one area where support from the nurse-in-charge, taking the family to one side to explain what is happening, or from a support worker, who would often get to know the family and provide continuity (FG4), made a positive difference to the nurse caring for the patient at the bedside. Communication processes between MDT members and between senior and junior nurses were important, in particular the role-modelling: ‘*good communication flows through to everyone*’ (FG6/Doctor). However, it was clear that nurse staffing affected the quality of communication between professions in situations like ward rounds; for example adequate staffing meant the nurse in charge could focus on discussions about the patient’s management, rather than being interrupted by staffing concerns (see Table 3).

The risks of admitting a patient when there were not enough nurses were balanced against the resource drain of transferring the patient to another hospital: ‘*pulling a doctor and a nurse out of the staffing for two or three hours*’ (FG7/Doctor). It was clear that allocation decisions were not based solely on nurses’ experience or education; confidence and trust in the nurses also had a key role to play. Tailoring allocations to staff wellbeing as well as patient need came through as key to getting these decisions right:

> ‘*you just wouldn’t put the nurse there* (with a particular type of patient) *for two consecutive days because it’s unfair on your nurse…. we are always very receptive to how people are – how the staff are feeling. And that does have a bearing on how you staff the unit*.’ (FG 6/Sister).

Decisions about staffing, mainly allocation and re-allocation throughout the shift, were emphasised: ‘i*t’s not that one allocation first thing in the morning. It’s how that allocation works through the day. It’s quite an organic thing maybe* (FG6/Sister). The acuity of the patient was the ‘*most important’* driver (FG6/Sister) but care needs, such as bowel care, pressure area care (FG3/nurse) and the need for family support (FG5/Matron) were also factored in where possible. By contrast, there was a clear perception that decision-makers from outside of critical care focused on levels of care and the nurse:patient ratio (structure factors) to make decisions, usually about moving ICU nurses to the ward to fill gaps. A network participant also reported a ‘*lack of understanding* (from managers outside of ICU) *of what it means to be a 24/7 service*’ (Int 8/Network Lead). The importance of avoiding ‘levels of care’ terminology when discussing staffing with managers outside of ICU was evident in one setting:

> P1: *that takes a bit of justification sometimes. But that’s why, on our unit, when we report back to the powers that be, we don’t refer to level 3, 2 or 1* (level of sickness). *We refer to patients that are one to one* (nurse:patient ratio), *one to two or ‘fit for the ward’. So, that they can be easily justified and they can understand why on this particular day we might be fully staffed but actually … we have no beds because of the acuity of the unit based on one to one, one to two or ward-fit patients…*..
>
> P3: …*and I think when you can’t achieve that and a nurse ends up looking after two people – the impact is actually massive I think*. (FG5/Matrons)

Decisions to move nursing staff sometimes happened within the ICU to balance out the workload for ‘*certain activities such as a patient admission*, [which] *need additional staffing*’ (Int 5/Network Lead), or between ICUs in the same hospital, including from general ICU to paediatric ICU. This latter staffing move ‘*causes upset’* (FG7/Staff Nurse) because it left staff feeling vulnerable ‘*it’s the fear of making an error… you’re terrified of getting it wrong*’ (FG7/Staff Nurse). As discussed above, staff were also moved from ICU to the wards either for a shift or, in one setting, for a longer secondment to the ward (FG7/Sister). This was also perceived as damaging for nurses: ‘*it breaks people* (to be constantly asked to work on the wards)’ (Int 4/network lead). There was also a perception that ICU nurses were seen as a ‘*feeding station for the wards*’ (Int 8/Network Lead), providing a bank of staff to fill gaps in ward staffing.

#### Impact on patient, staff and ICU flow outcomes

Participants identified a number of outcomes that they perceived to be influenced by nurse staffing but also identified the challenges of demonstrating some of these outcomes in the data currently collected. Categories in this theme were: adverse events; staff support; ICU flow and nursing workforce (see Table 4).

**Table 4.** Data excerpts related to *Impact on patient, staff and ICU flow outcomes* theme.

| **Outcomes** |  |
| --- | --- |
| Adverse events | *Increased mortality... increased morbidity but that takes a longer time to play through. Some of it you can see within the hospital stay – so a catheter ulceration. Or a pressure ulcer. Or UTIs. Staff – patient experience Also patient flow: are we having to do more transfers? Have we had to close beds? Have we had to cancel any theatre lists through lack of ICU beds? (Int 1/Policy Maker)*<br>*The two big red flags for me there are people leaving and a high number of cancellations on training courses, such as ITU transfer training, and it's always the off-duty cited as the reasons (Int 7/Network Lead)*<br>*The literature shows that getting a sick patient out of bed, or at least getting them moving in bed... it reduces length of stay and halves mortality.....but we just can't do it consistently [because of lack of staffing] FG1/Staff Nurse* |
| Staff support | *We had a situation one time that we lost three patients in the same day. In two to three hours. And the nurse in-charge on that day, she was brilliant because she gathered all the nurses that were looking after the patients and the healthcare supporters. Even the students. We just sat in the office and had a discussion. A debrief about how we felt. (FG4/staff nurse)*<br>*I allocated [a junior nurse] to a lower acuity patient at the start of the day. Absolutely stable. Of course within an hour of going into the shift [the patient] became acutely unwell and I said, 'Take this the right way. But I'm going to re-allocate you because he's become very, very unwell and I don't want to... 'I said, 'I don't want to break you.' And the patient was then safe and the nurse was safe. (FG5/Sister)* |
| ICU flow | *We have quite significant vacancies in the larger units; that doesn't mean that you dilute the nurse:patient ratios. It may mean that we have beds shut because of that ... they stick to the ratios but it has an impact on the service. (Int 9/Network Lead)*<br>*Where we lack the data is the patients that don't need to come in immediately but we don't have the staffing. It's very difficult for a clinician to declare that in the [patient's] notes that 'this patient should be in intensive care right now'. Erm. What they don't want to do is stand up in a court of law and have to defend why that patient wasn't. So they'll put* |
|  | *something like, 'Conversation with critical care. To manage the patient on the ward at the moment with critical care support.' which means we haven't got a staffed bed and we'd move them if we could (Int 10/Network Lead)* |
| Nursing workforce | *We've seen, in specialist areas, an exodus of our really good senior Band 5s – upper Band 5s – they leave to go into advanced practice roles [outside of ICU]. (FG4/matron)*<br>*It's the fear of making an error and making an error for a baby – you know – it can be fatal and even just getting a blood gas – getting a bit of air in – you're just terrified, you know, of getting it wrong. (FG7/staff nurse)*<br>*You are looking at the more measurable factors initially. Such as vacancies, sickness, recruitment and retention. And then that makes you ask questions about morale, culture, where is the unit [in relation to those issues]? (Int1/Policy maker)* |

Most of the outcomes discussed were negative outcomes for patients or staff; safe staffing was most commonly described in patient safety terms. Network participants identified a number of outcomes that they see as ‘red flags’ (potential risks to safety), particularly adverse events such as nasogastric tube displacements (Int 4/Network Lead), and pressure injuries (Int 1/Policy Lead). Focus Group participants emphasised the lack of time to provide full care for the patient, such as brushing hair and teeth and planning for the next stage in the patient’s care, and the lack of time for nurses to take breaks and remain hydrated (FG3/Staff Nurses and Sisters). Doctors also articulated an impact on medical workload of having experienced ICU nurses, for example in prioritising the actions needed after the ward round (FG1/Doctor; FG7/Doctor), but in a more general way they also emphasised ‘*the importance of the art and skill of the senior nurse’* (FG6/doctor).

Having enough staff to provide support to colleagues during tough shifts was deemed important. Examples included the reallocation of experienced nurses to work closely with, and support, less experienced staff (FG4/Staff Nurse) and the matching of nurse experience with patient level of care so that the situation was safe for patients and staff (FG5/Staff Nurse). Receiving good feedback from the nurse in charge and/or colleagues was reported to make a big difference for team morale and job satisfaction (FG 4/Staff Nurse and Sister). Important outcomes for nurses included having the time to provide fundamental care such as hair washing and nail care.

Patient flow outcomes such as delayed admission (FG 1/Staff Nurse), transfers for non-clinical reasons (Int 1/Policy Lead) and newly closed beds were also important outcomes suggesting to participants that nurse staffing may be inadequate. Weaning from sedation medication is important in preparing patients for ICU discharge but often delayed because of nurse staffing: ‘w*e sometimes sedate patients for longer if there aren’t enough staff’* (FG 7/Doctor). Patient rehabilitation, usually led by physiotherapists, was also delayed, especially when the nurse was managing two patients and not able to assist physiotherapists (FG7/Physio). Other aspects of physiotherapy treatment were also affected by nurse staffing: ‘*we want to go in and bag* [manually ventilate to loosen secretions] *the patient but they’re often cardiovascularly unstable so we need a nurse to be there in case they need a bolus* [of medication] *or something…. Often we can’t get on with what we need to do* (FG4/Physio).

Nursing workforce outcomes discussed by participants included retention of nurses, nurse vacancies, use of agency staff (Int 9/Network Lead), loss of education opportunities (FG1/Staff Nurse) and the number of shifts without supernumerary staff (Int10/Network Lead). The risk of allocating two high dependency patients to one nurse was also echoed across the interviews and Focus Groups: ‘*when one level 2 patient goes off* (deteriorates) *another level 2 patient is left unattended* (Int 7/Network Lead). The impact of being allocated two level 2 patients was articulated by some nurses as inducing a feeling of failure because they could not spend enough time with either patient (FG7/ Staff Nurse).

The challenges of demonstrating outcomes, particularly of reduced staffing, were acknowledged:

> ‘*the patient case-mix programme measures things like length of stay and mortality but these are not qualitative enough for nursing workload, but there are flaws in subjective data too, as things like patients and family satisfaction can be skewed by gratitude* [for surviving the ICU stay]’ (Int 8/Policy Lead).

#### Interaction between the themes

The interaction between the three themes is depicted at Figure 2; decisions about staffing were made throughout a shift and were influenced by a combination of factors illuminated in the three themes. Participants often described structural, process-related and outcome factors that influenced staffing decisions. An example was narrated in one Focus Group, comprised mainly of Matrons, where decisions about staffing depended on the *structure* of the MDT staffing for that shift, whether workaround *processes* were feasible and the *outcome* of current staffing, with regards to patient rehabilitation (FG5). Participants described reallocating staff during a shift, where possible, if patients were not going to get the care they required. Similarly, the complex interplay between the structure, process and outcome factors could constrain or enable decisions about nurse staffing.

**Figure 2.**
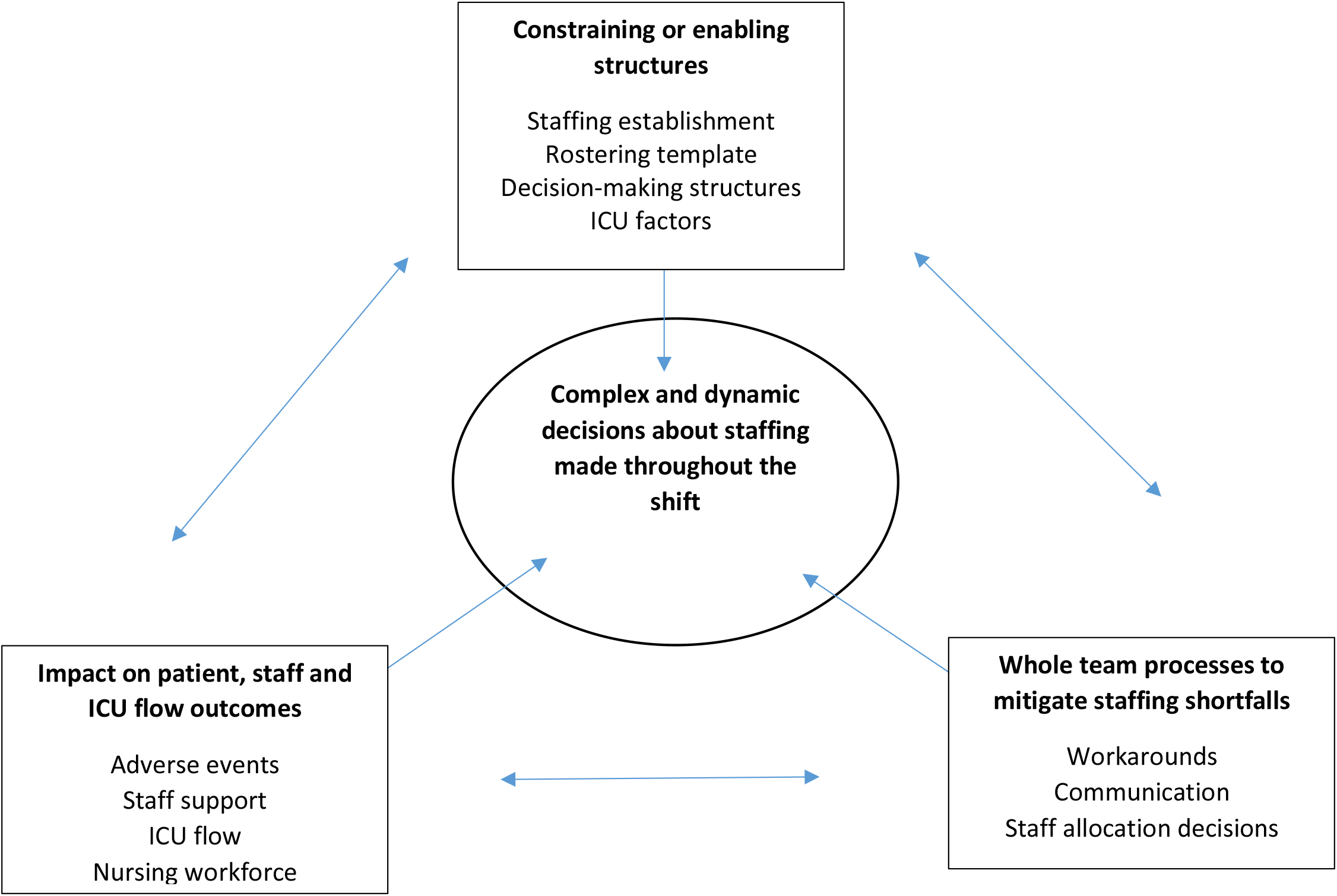
Managing nurse staffing: interaction between themes.

## Discussion

Previous studies provide evidence that outcomes such as patient mortality and increased risk of nosocomial infection are associated with nurse staffing in ICU (Rae et al. 2021). Analysis of our qualitative data from focus groups and individual interviews with 66 healthcare professionals showed three key factors influencing the complex and dynamic organisation of nurse staffing: multi-professional interdependence and workarounds; ICU geography; and staff wellbeing.

The ICU care of critically ill patients is typically delivered using inter-professional care bundles, for example the ABCDE bundle for managing sedation, delirium and early mobility (Pun et al. 2019) and the ventilator associated pneumonia (VAP) reduction bundle (Rello et al. 2013), with clear responsibilities for the different professions but, ideally at least, shared mental models (Botley et al. 2019, Liu et al. 2021). Hence the blurring of roles evidenced in our findings should not be a surprise. However, the examples, including workarounds, provided by our participants were not part of a planned inter-professional care bundle and carried an implicit assumption that they will understand each other’s roles, e.g. that the physiotherapist will know whether s/he needs to alert the RN to the observations just recorded. There is also an implicit assumption that interdisciplinary working will take place; our participants indicate that patient safety relies on this. However, there is no infrastructure to support this, for example knowledge/sharing of rosters between professions or joint planning, with evidence that hierarchical issues and cognitive biases persist (Liu et al. 2021).

Staffing guidance assumes that healthcare professionals work in silos, rather than in the multi-professional models portrayed by some of our participants and reflected the work cited above (Pun et al. 2019, Rello et al. 2013, Botley et al. 2019). The distinct contribution of individual professions to the management of the critically ill patient has been articulated, particularly in the many studies evaluating care bundles (Pun et al. 2019, Rello et al. 2013, Botley et al. 2019); our findings do not contradict this. However, there is unlikely to be a one size fits all solution and the potential opportunities for different staffing models are likely to be highly dependent on other professions.

Hence, any change to staffing models needs to take into account how different professions work together, and to formalise this in staff planning processes.

Processes are more amenable to change than structures (Mountford & Shojania 2012), which may explain why workarounds were quite dominant in the Focus Group discussions. The number of processes, including decision-making, reported by our participants to make the ICUs ‘safe’ for patients and staff highlight the importance of undertaking process evaluation alongside implementation of a change to staffing models. Process evaluation, as a component of a research study, captures the context in which changes are introduced and other factors that might illustrate why change does or does not become embedded.

The impact of the workplace layout on nurse workload has been reported in previous studies. The increase in nurses’ workload of changing from multi-occupancy to single room layout, has been reported in hospital wide and ICU settings (Maquire et al 2013, Lin et al. 2016). In Lin et al’s (2016) exploration of an ICU relocation, participants voiced concerns about patient and staff safety with the move to a single room model, whilst Leon-Villapalos et al (2020) found that perceptions of safety in ICU were negatively correlated with higher patient numbers and higher percentages of patients managed in single rooms. These studies, together with our findings, suggest that patient safety in ICU may not be best served by blanket ‘ratios’ approaches to nurse staffing, intended to apply uniformly across health services.

Staff wellbeing has arisen as a major concern during the COVID-19 pandemic, with one study in nine UK hospitals showing evidence of 49% of ICU nurses meeting criteria for post-traumatic stress disorder (168/344) and/or depression (167/344) (Greenberg et al 2021). Similar levels are reported in other studies (Rattray et al 2021, Sampaio et al 2021). Whilst these findings are important for future recruitment and retention of ICU nurses, our findings show that, among our participants, the shift-by-shift organisation of nurse staffing takes account of staff wellbeing were possible. Our Focus Group interviews were conducted before the COVID-19 pandemic, demonstrating that concerns about staff wellbeing are not purely pandemic-related.

Across the interviews and Focus Groups it was evident that a constant re-visiting of decision-making about staffing happened throughout the shift; this is reflected in the categories for structure, process and outcome themes. Most of the outcomes described are unintended consequences of the staffing model, or decisions in relation to the model. For example, the review of the number of level 2 and level 3 patients against the number of RNs on a shift, by a decision maker with limited understanding of ICU patient needs, might lead to an ICU RN being deployed to a ward to cover staffing shortages. However, the ICU shift leader cannot move the ICU RN back to ICU when a new patient is admitted (unintended outcome). Some of these unintended outcomes warrant exploration in empirical studies designed to examine staffing models.

### Strengths and Limitations

This national study recruited participants from ICUs of different size, with different heath service configurations from across England. The inclusion of staff from different professions has provided a more complete picture of factors influencing, and influenced by, nurse staffing in intensive care units. We conducted the focus groups before the pandemic, and the individual interview participants reflected on the organisation of staffing pre-pandemic, hence it is not known the extent to which perceptions about nurse staffing may have changed. However, our findings reveal a picture of under-staffing, at times, across all settings; this situation will not have improved during the pandemic.

There was a, sometimes implicit, focus on patient and staff safety across the interviews, hence Donabedian’s model was a useful framework with which to structure the initial analysis. The Donabedian approach did not detract from inductive generation of themes, but provided an overarching framework for analysis, facilitating constant comparison across the data sources.

### Conclusions

Whilst ratios were clearly used to set the nursing establishment, it was clear that rostering and allocation/re-allocation during a shift took into account many other factors such as staff wellbeing, the nursing needs of patients and family members, the ICU layout and the experience of all members of the multi-professional team. This has important implications for the future planning for ICU nurse staffing and highlights important factors to be accounted for in future research studies.

The findings have the potential to feed into discussions about the national tariff for critical care and quality metrics to be included in commissioning contracts.

## Data Availability

All data produced in the present work are contained in the manuscript.

## Acknowledgements

The SEISMIC research team comprises: Suzanne Bench, Fiona Boyd, Carole Boulanger, Chiara Dall’Ora, Ruth Endacott (CI), Jenny Gordon, Doug Gould, Peter Griffiths, David Harrison, Jeremy Jones, Thomas Monks, Paul Mouncey, Natalie Pattison, Susie Pearce, Annette Richardson, Kathy Rowan

